# A flow self-regulating STA-MCA bypass based on side to side fashion anastomosis for adult patients with moyamoya disease

**DOI:** 10.1101/2022.06.05.22275949

**Authors:** Jianjian Zhang, Miki Fujimura, Tsz Yeung Lau, Jincao Chen

**Affiliations:** Department of Neurosurgery, Zhongnan Hospital of Wuhan University, Wuhan 430071, China; Department of Neurosurgery, Hokkaido University Graduate School of Medicine, Sapporo, Japan; Department of Neurosurgery, Houston Methodist Hospital, Houston, TX, USA

**Keywords:** Direct bypass, End-to-side bypass, Side-to-side bypass, Moyamoya disease, Adult

## Abstract

**OBJECTIVE:** Side to side (S-S) fashion superficial temporal artery-middle cerebral artery (STA-MCA) bypass was reported for treating a special moyamoya disease (MMD) patient with collaterals arising from the donor STA. However, the S-S technique is not routinely performed to date and its benefits are still unknown for adult MMD. The purpose of this study is to investigate the possibility of routine use of the S-S technique for adult MMD.

**METHODS:** The authors retrospectively analyzed the clinical data from 50 adult patients (65 hemispheres, including 30 in end to side [E-S] group and 35 in S-S group) with MMD underwent STA-MCA bypass. The patients’ demographics, clinical courses, technical details, intraoperative blood flow, post- and preoperative relative cerebral blood flow (rCBF) values, modified Rankin Scale (mRS) scores and short-term revascularization results were compared between the two groups.

**RESULTS:** There was no significant difference observed in terms of baseline characteristics, bypass patency rates, post-/ preoperative rCBF values, incidence of cerebral hyperperfusion syndrome (CHS), improvement of mRS scores and short-term revascularization results between the two groups (P all > 0.05). Intraoperative blood flow analysis showed the increase of STA flow in the E-S group was significantly higher than that of proximal STA flow in the S-S group (P = 0.008 <0.05). Although the increases of proximal and distal recipient flow in the E-S group seemed higher than those in the S-S group, the results were not statistically significant (P = 0.086 in proximal and P = 0.076 in distal). The CHS symptoms were milder and their duration time was much shorter in the S-S group. The follow-up angiographic data of the representative case amazingly demonstrated that all the frontal and parietal STA branch and occipital artery participated in the postoperative collateralization.

**CONCLUSIONS:** S-S anastomosis can achieve comparable clinical effects to standard E-S construction. S-S anastomosis used in adult MMD demonstrated mild CHS symptoms with short duration time and had the potential to arouse all scalp arteries as donor sources for revascularization through the intact distal STA by a flow self-regulating fashion.

## Introduction

Direct or combined revascularizations for the purpose of increasing intracranial blood flow are considered to be effective in the treatment of moyamoya disease (MMD).^1^ However, postoperative complications, such as cerebral hyperperfusion syndrome (CHS), infarction and hemorrhage, have always been the focus of debate in MMD clinical practice. ^2,3^ Among these surgical procedures, the end-to-side (E-S) superficial temporal artery-middle cerebral artery (STA-MCA) anastomosis has been the most widely accepted technique. ^2^

In 2019, Lawton et al. performed a unconventional side-to-side (S-S) STA-MCA bypass surgery on one MMD patient in order to protect the developed native pial collaterals arising from the parietal STA branch which planned as the donor artery.^4^ Inspired by this study, we attempted to apply this S-S technique as a routine treatment for patients with adult MMD.

In this study, we performed the novel S-S STA-MCA bypass surgery on 35 adult MMD hemispheres, and retrospectively analyzed and compared the detailed clinical, angiographic, and intraoperative blood flow measurement data to those of 30 adult MMD surgical hemispheres who underwent E-S bypass just at almost the same period. We further describe the implementation details of this technology and discuss the possibility of routine use of the S-S technique for adult MMD.

## Methods

### Study Design and Patient Population

In this study, we included all adult patients with MMD who recevied a STA-MCA anastomosis combined with encephaloduromyosynangiosis (EDMS) revascularization procedure in our hospital between June 2021 to December 2021. The diagnosis for each patient was established by digital subtraction angiography (DSA). All patients satisfied the diagnostic criteria of the Research Committee on Spontaneous Occlusion of the Circle of Willis of the Ministry of Health, Labor, and Welfare, Japan.^5-7^ The indication for surgery included patients with symptomatic MMD (ischemic or hemorrhagic) with apparent hemodynamic compromise observed by single-photon emission computed tomography (SPECT) and a modified Rankin Scale (mRS) score less than 2. This study protocol was approved by the Institutional Review Board at Zhongnan Hospital of Wuhan University (approval number: Kelun-2020063) and was in accordance with the Declaration of Helsinki revised in 1983. Written informed consent was obtained from all patients.

### Surgical Procedures

The surgical procedure of E-S fashion STA-MCA anastomosis was almost the same as those already introduced by previous articles and books. ^8-10^

Our S-S fashion STA-MCA bypass procedure for adult MMD is as following and its diagram is shown in Figure 1: The patient is placed in supine position with the head rotated to contralateral side. Using Doppler ultrasonography, the courses of the frontal and the parietal STA branch are identified. Along the course of the selected STA branch, it is meticulously dissected from the surrounding tissues. Subsequently, the temporalis muscle is incised to the bone along the root of the temporal-parietal skin flap and elevated anteriorly to expose a 7×8 cm area of temporal and frontal bone junction, allowing for exposure of the parasylvian cortical arteries (PSCAs). A heart shaped craniotomy is performed to preserve the MMA. After cortical surface exposed, an indocyanine green videoangiography (ICG-VA) is performed to help selecting recipient artery. The number of available recipient arteries of each patient was recorded. Interrupted suturing technique was used for anastomosis. Most of time, 3-4 stitches for each wall are enough because of the typical small arteriotomy in our procedure. After direct anastomosis completed, an ICG-VA is performed to confirm patency of the bypass.

**Figure 1.**
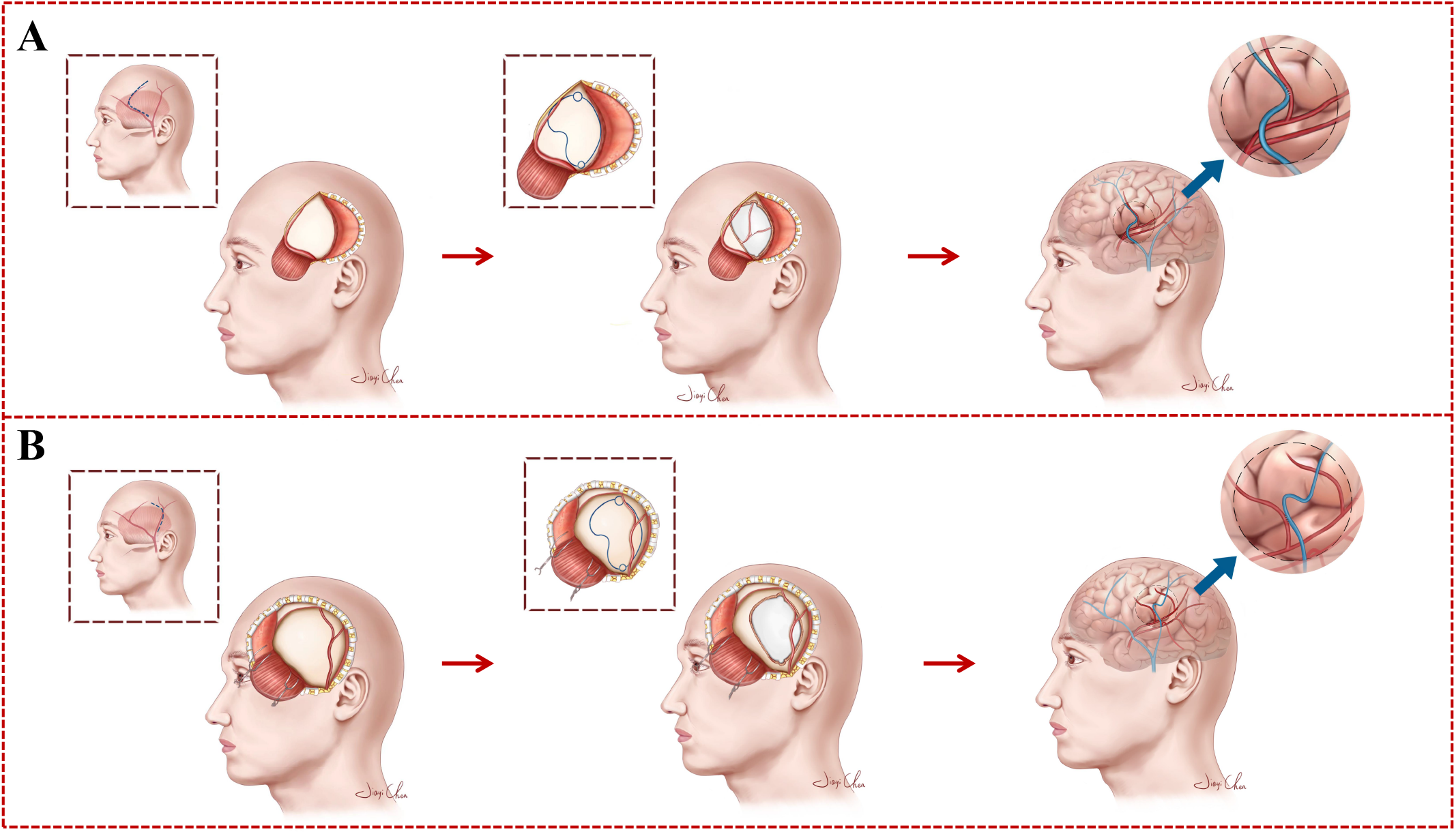
Diagram of surgical procedures of the novel side to side STA-MCA anastomosis for adult MMD (skin incision, temporal muscle dissection, craniotomy, and direct anastomosis). **A**. the frontal STA branch is selected as the donor artery. **B**. the parietal STA branch is selected as the donor artery. In our cohorts, the parietal STA branch is more commonly selected as the donor artery though its length is limited because the frontal branch frequently runs out of the hairline.

### Pre- and postoperative measurement of rCBF from SPECT

All the patients underwent technetium-99m ethyl cysteinate dimer (^99^mTc-ECD) brain SPECT imaging. Postprocessing of raw SPECT source images was completed centrally on a dedicated workstation (Discovery NM/CT 670 Pro, GE). Preoperative CBF was measured within 1 week before surgery and postoperative CBF was measured 1 day after surgery in all the patients. Anastomosis sites were recorded during surgeries and then confirmed by the original axial slice of postoperative MR angiography. Standardized regions of interest (ROIs) were drawn manually as a circle with a diameter of 1 cm in the cortical areas at the site of the anastomosis and the ipsilateral cerebellum. The relative cerebral blood flow (rCBF) value was defined as the ratio between the CBF value of the anastomosis site and that of the control region in the cerebellum. The blood flow change at the anastomosis sites was demonstrated by the ratio of post- / preoperative rCBF. ^11^

### Intraoperative Blood Flow Analysis

Pre- and post-anastomosis ICG-VA was performed with a commercially available operating microscope (OPMI Kinevo 900, Carl Zeiss). A body weight-adapted dose of 0.25 mg/kg ICG (Pulsion Medical Systems) was injected through a central venous catheter followed by a 10-ml bolus of sterile saline. After obtaining the color-coded FLOW 800 blood flow map which automatically generated by the microscope, predefined square ROIs were manually placed into the chosen recipient arteries in all patients, both proximal and distal to the anastomosis site (Figure 2A and 2B). While there was only one ROI placed into the donor STA in the E-S group, there were 2 ROIs placed in both proximal and distal to the anastomosis site in the donor STA in the S-S group. In both groups, another non-recipient PSCA with the highest velocity was selected as the control artery from the pre-anastomosis FLOW 800 blood flow image. After all the ROIs placed, several parameters including delay time (s), velocity (/s), time to peak (s) and rise time (s) were automatically generated. Due to lack of references in calculating semiquantitative flow from Kinevo 900, relative donor and recipient flow was first normalized and recorded by the following method in this study: the velocity values of the donor and recipient arteries / the velocity value of the control artery. The blood flow change in the arteries resulting from the bypass was demonstrated by the ratio of post- / pre-anastomosis relative flow. Distal STA flow in the S-S group was special and absent in the E-S group. Except for comparing the changes of proximal and distal recipient flow between the two groups, we also analyzed the difference between the changes of STA flow in the E-S group and those of proximal STA flow in the S-S group (Figure 2C and 2D).

**Figure 2.**
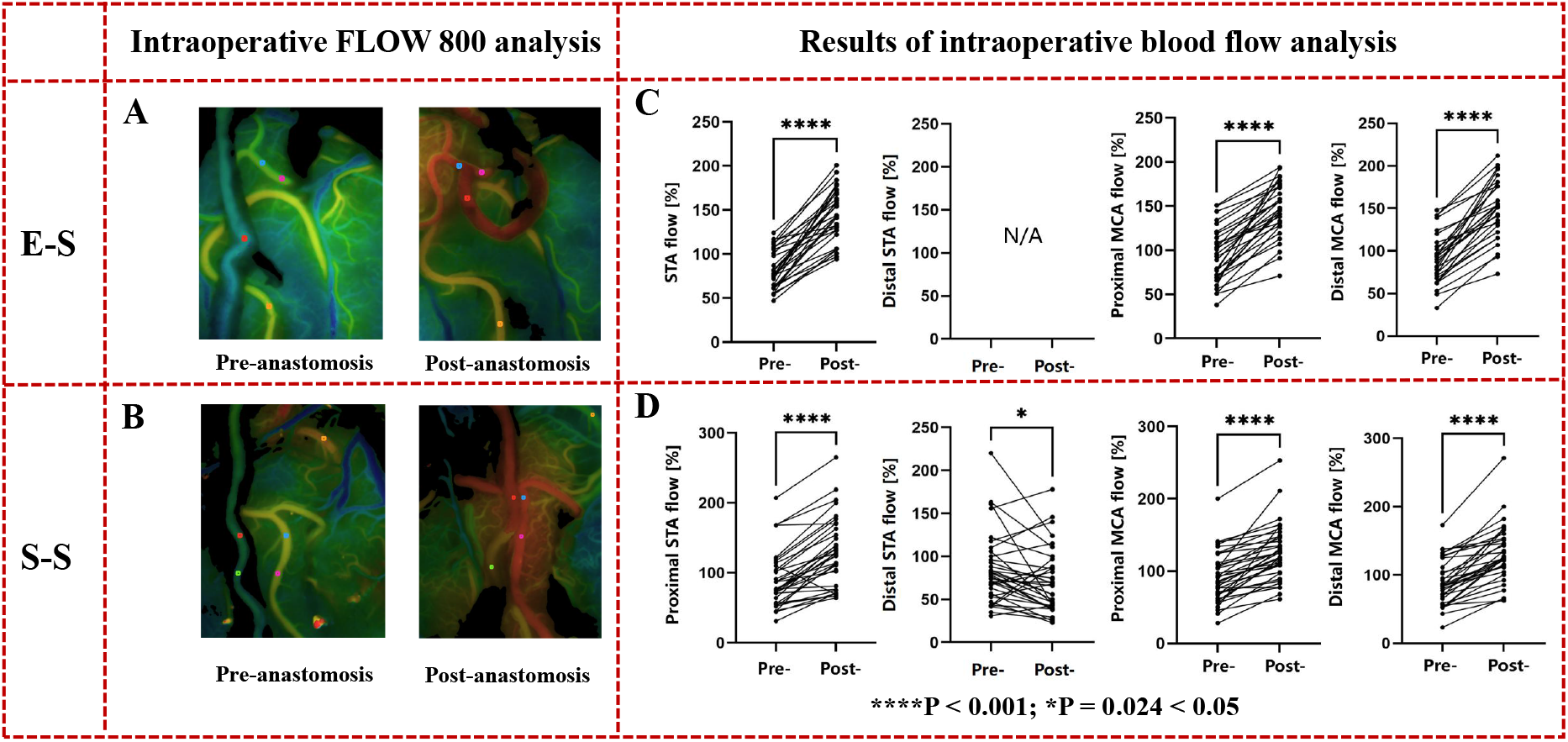
Illustration of intraoperative blood flow analysis by FLOW 800 ICG angiography in Kinevo 900. **A and B** illustrate the intraoperative blood flow analysis procedures in both groups. **A:** E-S group, **B:** S-S group. Predefined square ROIs are manually placed into the chosen recipient arteries, both proximal (blue) and distal (purple) to the anastomosis site. There is only one ROI is placed into the donor STA (red) in the E-S group but 2 ROIs are respectively placed in proximal (red) and distal (green in Figure 2B) to the anastomosis site in the donor STA in the S-S group. Another remote non-recipient PSCA (yellow) with the highest velocity is selected as the control artery. The post-/pre-anastomosis blood flow changes are calculated. **C and D** demonstrate the results of intraoperative blood flow analysis. Proximal STA flow in the S-S group, STA flow in the E-S group, proximal and distal recipient flow in both groups all significantly increased after anastomosis. Distal STA flow which can only be observed in the S-S group is special and interesting. There was a significant difference between post- and pre-anastomosis distal STA flow in the S-S group. ****P < 0.001; *P = 0.024 < 0.05.

### Comparison of clinical courses between the two groups

The following parameters including numbers of available recipient arteries, temporal occlusion time during anastomosis, incidence of postoperative CHS, duration time of CHS symptoms, bypass patency and revascularization results at short-term follow-up (3 months) were compared between the two groups.

The perfusion status and patency of anastomosis were detected by SPECT and MRA at days 1 and 2 after surgery, respectively. CHS was determined using the criteria in one of our previous studies.^12^ If CHS was confirmed for the patient, strict blood pressure control and edaravone administration were scheduled. Duration time of CHS symptoms in each CHS patient was recorded.

The DSA images at short-term follow-up (3 months) were analyzed for evaluating bypass patency and revascularization effect. The revascularization results were obtained by using the established Matsushima grading system^13^: grade A, filling of more than two-thirds of the MCA circulation; grade B, between one- and two-thirds; and grade C, filling of less than one-third.

### Statistical Analysis

Descriptive summary statistics are presented as mean ± standard deviation. Numerical data were compared between two groups using independent t-test and within the same group using paired t-test. Categorical variables were analyzed in contingency tables with Chi-square test and Fischer’s exact test. All analyses were performed with IBM SPSS Statistics Desktop, version 22.0 (IBM Corp.). The results with values of *P* < 0.05 were considered significant.

## Results

### Grouping and basic characteristics

Fifty adult patients with MMD (patients aged 28-69 years old, mean 49.52 years old) were included in this study. A total of 65 symptomatic hemispheres of these patients were performed direct STA-MCA (M4) anastomosis combined indirect EDMS. Consequently, the earlier 30 hemispheres that received E-S fashion anastomosis were included in the E-S group while the later 35 hemispheres performed S-S fashion anastomosis were included in the S-S group. In these cohort cases, 15 patients were bilaterally operated (the first using E-S and second using S-S). Only one hemisphere which was supposed to receive S-S anastomosis but finally changed to E-S anastomosis due to lack of available recipient artery.

The basic characteristics of enrolled patients as well as hemispheres were separately summarized in Table 1. There was no significant difference observed in terms of age, sex, initial onset type, Suzuki stage or surgical side between the two groups (*P* > 0.05 for those variables).

**Table 1.** Comparison of basic characteristics, technical details, radiological results between the two groups

|  | End to side<br>(n=30) | Side to side<br>(n=35) | P value |
| --- | --- | --- | --- |
| Age, Year | 48.23 ± 9.14 | 48.57 ± 10.81 | 0.893 |
| Sex, male (%) | 16 (53.3) | 17 (48.6) | 0.702 |
| Onset, n(%) |  |  | 0.223 |
| Ischemic | 20 (66.7) | 28 (80.0) |  |
| Hemorrhagic | 10 (33.3) | 7 (20.0) |  |
| Suzuki stage, n(%) |  |  | 0.664 |
| II | 3 (10.0) | 2 (5.7) |  |
| III | 12 (40.0) | 13 (37.1) |  |
| IV | 8 (26.7) | 14 (40.0) |  |
| V | 7 (23.3) | 6 (17.1) |  |
| Surgical side, n(%) |  |  | 0.786 |
| Left | 17 (56.7) | 21 (60.0) |  |
| Right | 13 (43.3) | 14 (40.0) |  |
| Donor artery (STA) |  |  | 0.600 |
| Frontal branch | 3 (10.0) | 5 (14.3) |  |
| Parietal branch | 27 (90.0) | 30 (85.7) |  |
| Numbers of available recipients | 2.83 ± 0.87 | 1.86 ± 0.73 | <0.001 |
| Temporary occlusion time (min) | 18.23 ± 3.79 | 23.34 ± 4.38 | <0.001 |
| Incidence of CHS | 3 (10.0) | 2 (5.7) | 0.857 |
| Post-/ pre- rCBF from SPECT | 1.198 ± 0.125 | 1.201 ± 0.134 | 0.921 |
| Post-/pre-anastomosis relative flow<br>from ICG-VA |  |  |  |
| Proximal STA flow | 1.831 ± 0.444 | 1.536 ± 0.430 | 0.008 |
| Proximal recipient flow | 1.698 ± 0.499 | 1.484 ± 0.485 | 0.086 |
| Distal recipient flow | 1.765 ± 0.443 | 1.567 ± 0.439 | 0.076 |
| Matsushima grading (3-month<br>follow-up, %) |  |  | 0.765 |
| A | 9 (34.6) | 10 (40.0) |  |
| B | 13 (50.0) | 10 (40.0) |  |
| C | 4 (15.4) | 5 (20.0) |  |
| N/A | 4 | 10 |  |
| Bypass occlusion (Follow-up, %) | 1 (3.3) | 1 (2.9) | 0.912 |

STA = superficial temporal artery; CHS = cerebral hyperperfusion syndrome; rCBF = relative cerebral blood flow; SPECT = single-photon emission computed tomography; ICG-VA = indocyanine green videoangiography; N/A= not available.

### Comparison of clinical characteristics between the two groups Technical details

1. Donor artery: There was no difference of donor branch selection between the two groups (*P* = 0.600). Although the STA frontal branch was more flexible than the parietal branch, much more parietal branches were selected as the donor arteries in both groups.
2. Recipient artery: In our center, we preferred to select a PSCA above the sylvian fissure as the recipient artery according to the symptoms and preoperative hemodynamic evaluation. The number of available recipient arteries in the E-S group (2.83 ± 0.87) was significantly more than that in the S-S group (1.86 ± 0.73) (*P* < 0.001).
3. Temporary occlusion time: The temporary occlusion time of PSCA in the S-S group (23.34 ± 4.38 minutes) was significantly longer than that in the E-S group (18.23 ± 3.79 minutes) (*P* < 0.001). The average difference was about 5 minutes.
4. Bypass patency rate: According to ICG-VA, the intraoperative bypass patency rates were 100% in both groups. Each group had only one graft occlusion found during follow-up. There was no significant difference of bypass patency rate between the two groups (*P* = 0.912).

### Characteristics of ICG-VA and intraoperative blood flow analysis

Proximal STA flow in the S-S group, STA flow in the E-S group, proximal and distal recipient flow in both groups are all significantly increased after anastomosis (P all < 0.001) (Figure 2). Although the increases of proximal and distal recipient flow in the E-S group seemed higher than those in the S-S group, the results were not statistically significant (P = 0.086 in proximal and P = 0.076 in distal). It was interesting that the increase of STA flow in the E-S group was significantly higher than that of proximal STA flow in the S-S group (P = 0.008) (Table 1).

Distal STA flow in ICG-VA could only be observed in the S-S group (Figure 2). Post-anastomosis distal STA flow was higher than pre-anastomosis distal STA flow in 10 hemispheres (10/35, 28.6%). However, most of time (25/35, 71.4%), distal STA flow significantly decreased after anastomosis. There was a significant difference between post- and pre-anastomosis distal STA flow (*P* = 0.024).

### Incidence of postoperative complications and duration time of postoperative CHS

The SPECT data showed there was no significant difference on the ratio of post-/pre-operative rCBF values between the E-S group (1.198 ± 0.125) and the S-S group (1.201± 0.134) (*P* = 0.921). Both groups had a relatively low incidence of postoperative CHS (5.7% in the S-S group and 10.0% in the E-S group, respectively) with no significant difference between the two groups (*P* = 0.857). However, it was interesting that the duration time of CHS symptoms of the two CHS patients (3 days, 6 hours, respectively) in the S-S group were much shorter than that of the three CHS patients (7, 9, 10 days, respectively) in the E-S group. Furthermore, we found the CHS symptoms in the S-S group were much milder than those in the E-S group. There was 1 postoperative cerebral hemorrhage and 2 seizures in the E-S group but none in the S-S group. (Table 2)

**Table 2.** Characteristics of the patients with perioperative complications

| No. | Sex | Age Range | Onset | Suzuki | Comorbidities | Method | Surgical side | Bypass site | TOT (minutes) | Post-/pre- anastomosis flow change |  |  |  | Post/pre SPECT | Complications | DT | mRS | MG (FU DSA) |
| --- | --- | --- | --- | --- | --- | --- | --- | --- | --- | --- | --- | --- | --- | --- | --- | --- | --- | --- |
|  |  |  |  |  |  |  |  |  |  | STA or P-STA | D-STA | P-recipient | D-recipient |  |  |  |  |  |
| 1 | F | 66-70 | Ischemic | 5 | Hypertension +Diabetes | E-S | Left | Frontal lobe | 28 | 1.240 | N/A | 1.205 | 1.204 | 1.039 | Basal ganglia small Infarction, hemiplegia | 1m | 2 | N/A |
| 2 | F | 36-40 | Ischemic | 4 | None | S-S | Left | Parietal lobe | 22 | 2.903 | 2.052 | 2.358 | 1.876 | 1.338 | Motor aphasia | 3D | 0 | N/A |
| 3 | M | 30-35 | Ischemic | 5 | None | S-S | Left | Frontal lobe | 21 | 2.194 | 2.263 | 3.256 | 2.437 | 1.441 | Sensory aphasia | 6h | 0 | B |
| 4 | F | 50-55 | Ischemic | 3 | Hypertension | E-S | Left | Frontal lobe | 18 | 2.386 | N/A | 2.232 | 2.481 | 1.304 | Motor aphasia, weakness of right limbs | 9D | 0 | B |
| 5 | M | 50-55 | Ischemic | 3 | None | E-S | Left | Frontal lobe | 15 | 1.669 | N/A | 1.625 | 1.742 | 1.495 | Small hematoma, motor aphasia, and seizure | 7D | 1 | A |
| 6 | F | 46-50 | Ischemic | 5 | None | E-S | Right | Frontal lobe | 19 | 1.485 | N/A | 1.722 | 1.705 | 1.359 | weakness of left limbs, seizure | 10D | 0 | B |

TOT = temporary occlusion time; P- and D-STA = proximal and distal superficial temporal artery; P- and D-recipient = proximal and distal segment of the recipient artery; SPECT = single photon emission computed tomography; DT = Duration time of the symptoms; mRS = modified Rankin Scale; MG = Matsushima grading; FU DSA = follow-up digital subtraction angiography; Sex (F and M) = female and male; E-S = end to side; S-S =side to side; N/A= not available.

### Clinical symptom improvement and mRS

The preoperative symptoms of most patients in both groups improved or remained unchanged after surgery at last follow-up. The postoperative mRS scores were significantly lower than preoperatively in both groups (*P* both < 0.001) (Figure 4). Only one patient in the E-S group experienced deterioration after operation. There were no significant differences of the preoperative or postoperative mRS scores between the two groups (*P* = 0.629 and 0.927, respectively). None of the patients experienced a new ischemic or hemorrhagic stroke since discharge.

**Figure 3.**
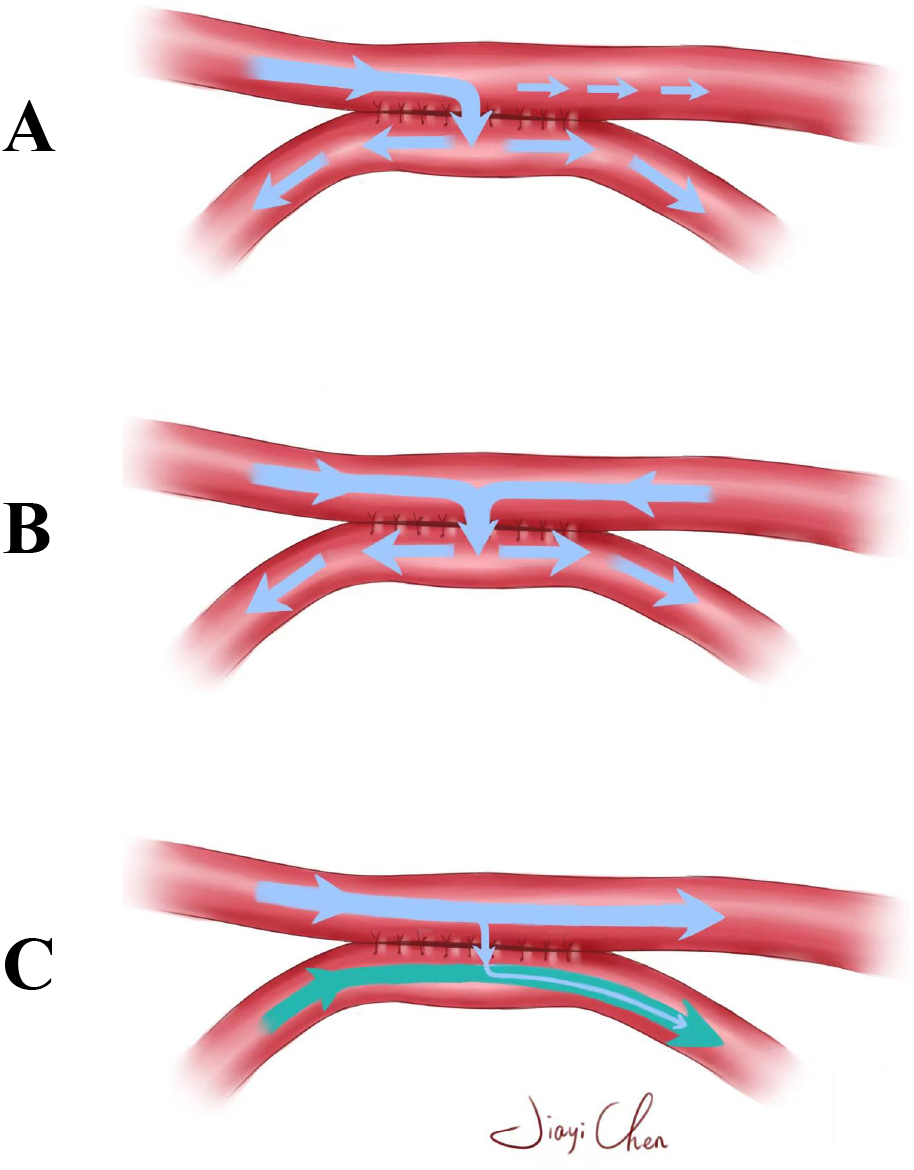
Typical post-anastomosis input and output blood flow patterns. **A**. S-S technique has the possibility to achieve the self-regulation of incoming blood flow through shunting extra blood flow into the distal STA based on the actual intracranial perfusion needs. **B**. S-S technique has the opportunity to attract the reserved distal STA flow to take part in postoperative cerebral revascularization through the anastomotic site if there is massive blood flow demand in the ischemic hemisphere. **C**. If the ischemic extent of the hemisphere is not very severe, most blood flow of the donor STA will automatically shunt into the distal segment and will not meddle unduly with preexisting intracranial blood flow.

**Figure 4.**
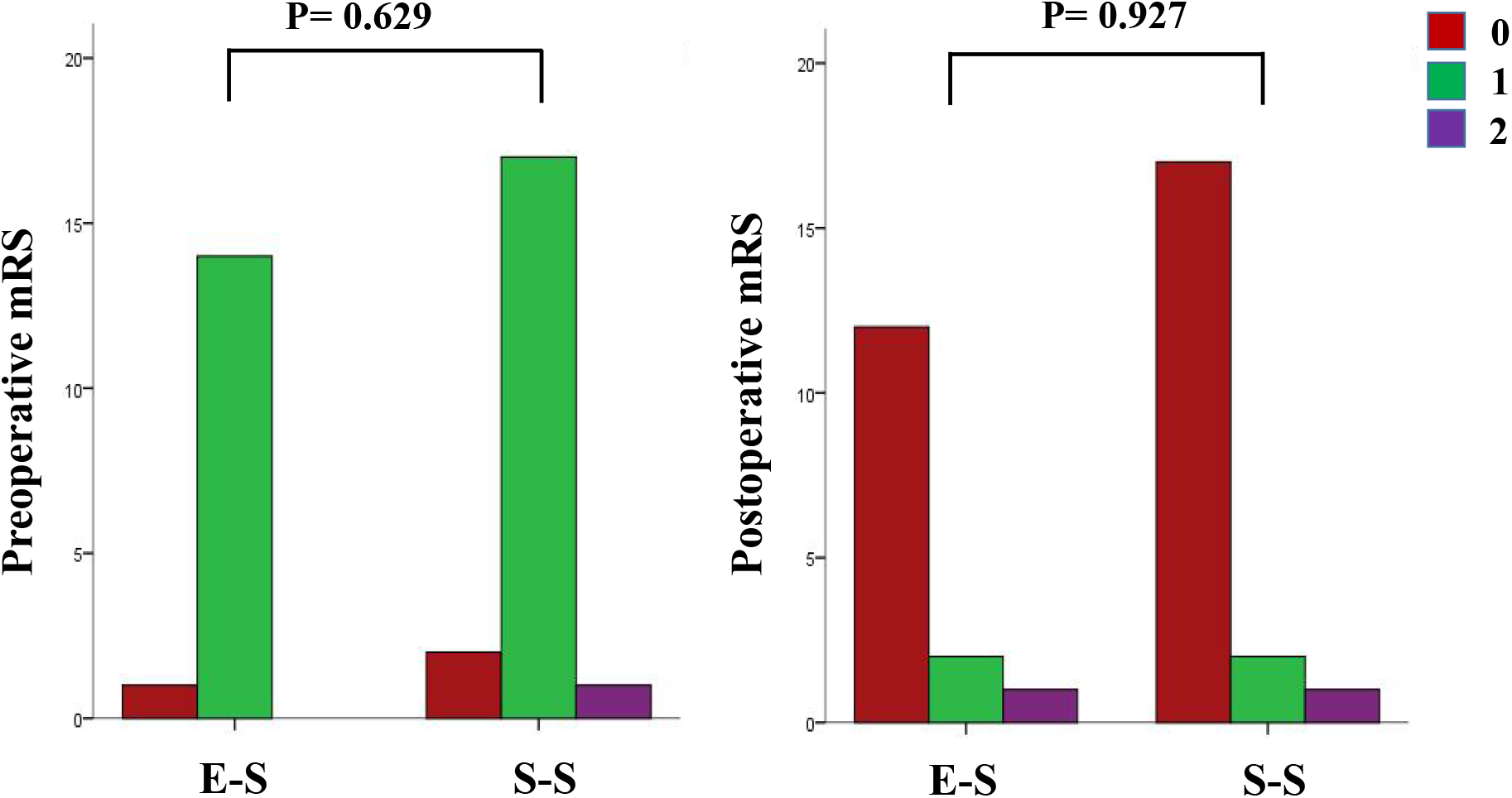
Graph of pre- and postoperative mRS scores in the two groups (excluding the 15 patients underwent bilateral surgeries) The mRS scores were significantly improved after surgery in both groups (P both < 0.001). There were no significant differences of the preoperative or postoperative mRS scores between the two groups (P = 0.629 and 0.927, respectively).

### Follow-up angiographic Outcomes

All the patients were suggested to perform a follow-up DSA examination at postoperative three months. However, there were only 26 (86.7%) cases in the E-S group and 25 (71.4%) cases in the S-S group ultimately accepted our suggestions. After careful analyzing of their preoperative and short-term follow-up DSA images, we found there was no significant difference of the revascularization results between the two groups (*P* = 0.765). Most of the patients in both the E-S group (84.6%) and the S-S group (80.0%) could achieve a relatively good outcome (Matsushima grade A or B). (Table 1)

### Representative case

This was a female patient (60-65 years old) who presenting with recurrent weakness of the right limbs for 1 month. DSA confirmed MMD (Figure 5A). We decided to perform the novel S-S anastomsis for her. The parietal STA branch was selected as the donor artery. According to the pre-anastomosis ICG image, there were 3 potential recipient arteries in the operating field. Intraoperative post- /pre-anastomosis blood flow changes in the proximal and distal STA, the proximal and distal recipient PSCA were 1.159, 1.096, 1.253, and 1.121, respectively (Figure 5B). The temporary occlusion time was 22 minutes. ICG showed the patency of the anastomosis. SPECT showed rCBF of the anastomotic site was significantly improved and the post-/preoperative rCBF value was 1.198 (Figure 5C). Postoperative blood pressure was maintained in the normal range, and she did not present any neurological deficit perioperatively. At 3-month short-term follow-up, she totally recovered from her preoperative symptoms and the mRS score was 0. Follow-up DSA showed a good angiographic outcome (Matsushima grade A and supplemental data) and typical distal STA flow and collateral changes after her S-S STA-MCA bypass (Figure 5A). This case demonstrated that S-S technique used in treatment for MMD had the potential to arouse all scalp arteries (the frontal and parietal STA branches and occipital artery in this case) as donor sources for revascularization through the intact distal STA by a flow self-regulating fashion.

**Figure 5.**
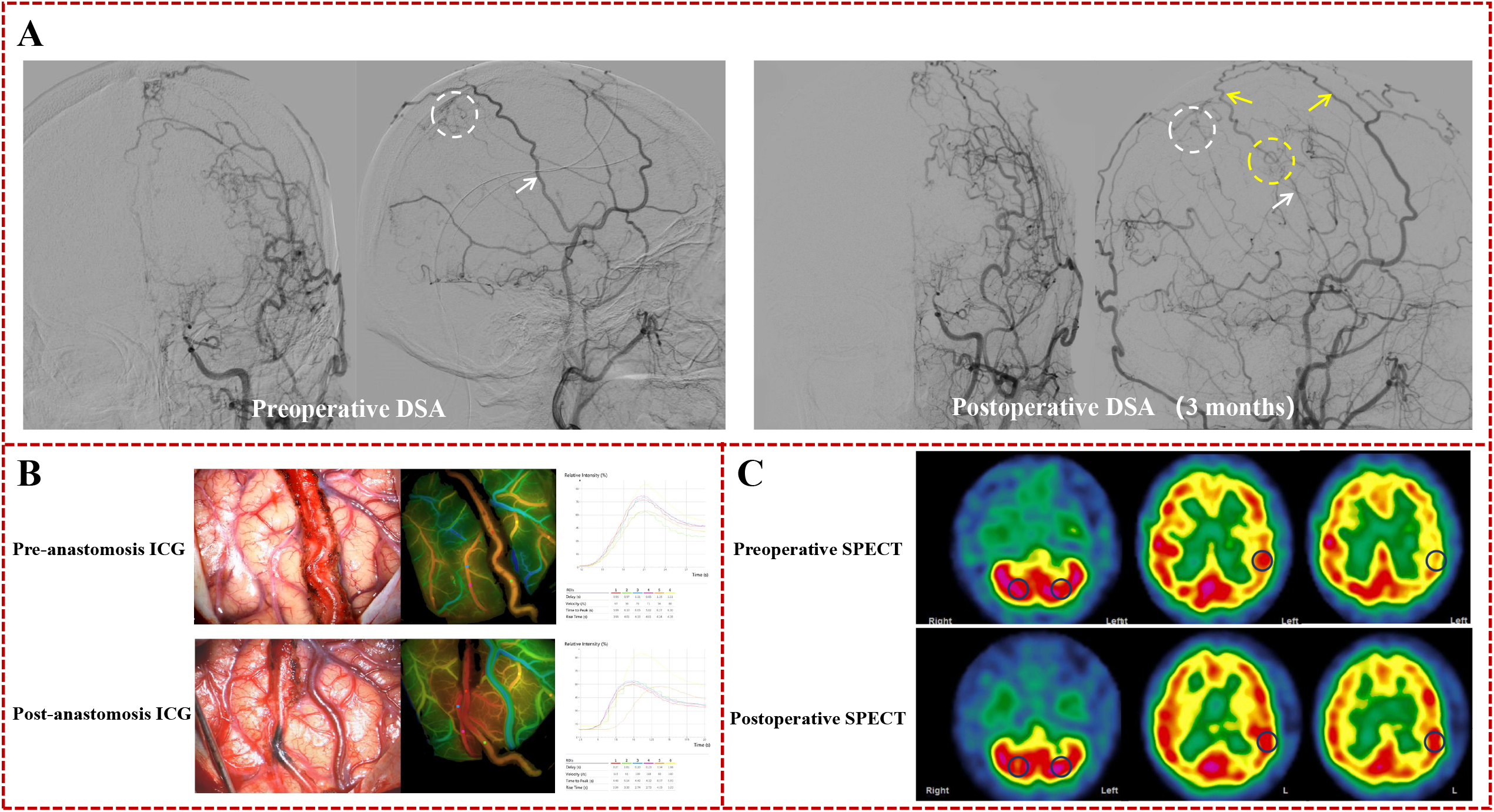
Representative case. This case was a female MMD patient who presenting with recurrent weakness of the right limbs for 1 month. **A**. Preoperative DSA (**left**) showed terminal occlusion of the left internal carotid artery. The Left common carotid artery (CCA) injection demonstrated collaterals to the paracentral lobule arising from the parietal STA (White circle). The parietal STA branch was selected as the donor artery (White arrow). 3-month follow-up DSA (**right**) showed interesting blood flow changes after her S-S STA-MCA bypass: first, the proximal segment of the parietal STA branch became much thinner but still keeping patent and providing blood flow into the brain (White arrow). Second, the distal segment of the frontal STA branch which not seen from the preoperative DSA image was observed (Yellow arrows) and very strong to provide blood flow into the brain through the anastomotic site (Yellow circle). Third, collaterals to the paracentral lobule was changed from the parietal STA to the left occipital artery after bypass (White circle). **B**. According to the pre-anastomosis ICG image, there were 3 potential recipient arteries in the operating field. Post-anastomosis ICG showed the patency of the anastomosis. **C**. SPECT showed rCBF of the anastomotic site was significantly improved and the post-/preoperative rCBF value was 1.198 (Blue circles showed the ROIs placed for calculating the rCBF values).

## Discussion

The technique of S-S STA-MCA bypass with maintaining distal outflow of the donor STA was previously described for treating a special MMD patient with collaterals arising from the donor STA.^4^ Our series is the current largest published series of this technique with an easy suturing method. The present study demonstrates the feasibility of routine use of this novel S-S anastomosis in treatment for adult MMD.

### Strengths of S-S STA-MCA bypass for adult MMD

#### (1) Technical available

E-S fashion anastomosis is now the most popular technique used in cerebral revascularization. It is familiar by most vasucular neurosurgeons and characterized by simple operation and high bypass patency. ^5^ To meet the various necessity of cerebral bypass, other techniques such as high-flow external-internal carotid (EC-IC) bypass using an interposition graft, intracranial reconstructions that reanastomose parent arteries, and S-S anastomosis are evolved.^4,14-16^ Although technically more complex than E-S anastomosis, the performance of these varieties is already proved to be generally favorable. Moreover, we did not use the intraluminal suturing techniques which specially designed for S-S anastomsis in this study.^4^ Instead, we performed a simpler S-S anastomosis using the suturing technique just like that in E-S anastomosis by flipping over the donor STA along its longitudinal axis (**Video, Supplemental Digital Content**). Of course, this technical modification can only be highlighted for a superficial S-S anastomosis, such as S-S fashion STA-M4 bypass. Consequently, due to the same suturing techniques, the bypass patency rates of the S-S group (intraoperative 100% and follow-up 97.1%) and E-S group (intraoperative 100% and follow-up 96.7%) were extremely similar in this study. Although the temporary occlusion time (average 5 minutes) in the S-S group is longer than that in E-S anastomosis, which partially represents the more complex of S-S anastomosis, this slight differences in temporary occlusion time is acceptable in such a superficial bypass for MMD.

#### (2) Self-regulating flow augmentation and few perioperative complications

Different from the traditional E-S bypass, the novel S-S bypass surgery has some special principles.

First, the biggest feature of S-S technique is that it preserves the distal outflow of the donor STA and has the possibility to achieve the self-regulation of incoming blood flow according to the actual intracranial perfusion needs through shunting extra blood flow into the distal STA (Figure 3A). The excessive incoming flow can quickly result in postoperative local hyperperfusion around the anastomosis and remote blood flow distribution disturbance, which led to CHS or remote cerebral ischemia at the same time by the “watershed shift” phenomenon.^17,18^ This uneven cerebral hemodynamic change confuses neurosurgeons in postoperative management of MMD patients. Once CHS or remote cerebral ischemia occurs, the management strategies are limited to strict blood pressure control, administration of minocycline, endaravone and so on.^19,20^ According to previously published studies, during this difficult period, the patient received E-S bypass can do nothing but passively wait for slow disappearance of neurological disturbance, and even suffers from more severe problems such as cerebral hemorrhage and infarction.^21^ However, in S-S bypass, the possible excessive blood flow can be shunted into the distal STA, so as to quickly relieve postoperative CHS and decrease the severity of relative symptoms. In the present study, although there was no significant difference of incidence of CHS between the E-S and S-S group (*P* > 0.05, may be due to small samples), the CHS symptoms in the S-S group were milder than those in the E-S group and had shorter duration time (Table 2).

Second, the S-S technique has the opportunity to attract the reserved distal STA flow to take part in postoperative cerebral revascularization through the anastomotic site if there is massive blood flow demand in the ischemic hemisphere (Figure 3B). From the 3-month angiographic images of the representative case, we observed that not only the donor parietal STA branch but also the frontal STA branch and occipital artery provided blood flow for her postoperative revascularization through the abundant scalp collateral anastomoses (Figure 5A). This situation can be vividly described as the neurosurgeons just open a door for the patients and left the rest to themselves. Based on a self-regulating fashion, the frontal STA branch self enlarged and became the major donor blood flow source of direct revascularization. The parietal STA branch which we planned as the donor artery became thin but still also provide blood flow to the brain through the anastomotic site.

Third, if the ischemic extent of the hemisphere is not very severe, most blood flow of the donor STA will automatically shunt into the distal segment and will not meddle unduly with preexisting intracranial blood flow (Figure 3C). Under this circumstance, we can regard the new S-S operation as an accelerated (encephalo-duro-arterio-synangiosis,EDAS) for treating MMD.

At last, preservation of distal STA can retain the developed intra- and extra-cranial compensatory collaterals and scalp circulation to the greatest extent, and “do no harm” to the patients while providing satisfactory effect of revascularization.

The main theoretical weakness is S-S anastomosis may lead to distal STA steal phenomenon. However, our FLOW 800 data demonstrated that although the post-/pre-anastomosis blood flow changes of the proximal donor STA in the S-S group were significantly lower than those of the donor STA in the E-S group, those of the recipient arteries had no significant difference between the two groups. Moreover, it is interesting that the post-/preoperative rCBF values from SPECT of the two groups also had no significant difference. These data shows S-S technique applied in MMD may result in at least comparable blood flow and perfusion improvement when compared to the traditional E-S technique.

Consequently, the above principles indicate the new S-S bypass can result in favorable revascularization with low risks of perioperative complications by a flow self-regulating fashion.

#### (3) Excellent clinical outcomes

The mRS scores of most patients were improved after surgery and the others remained the same to preoperative mRS except one patient in the E-S group who experienced perioperative cerebral infarction and hemiplegia. This result was consistent in previously published studies.^22,23^ The short-term follow-up DSA results showed that there was no significant difference in Matsushima grades between the two groups. The symptoms of most patients relieved or remained unchanged after surgery in both groups. Based on these results, we concluded that S-S anastomosis can achieve analogous clinical outcomes to traditional E-S anastomosis when used in treatment for MMD.

Taken together, we conclude the possible advantages and disadvantages of the two STA-MCA bypass techniques in Table 3. However, larger samples studies are needed to verify the ideas.

**Table 3.** Advantages and disadvantages of the two STA-MCA bypass techniques for adult MMD

| Variable | Traditional end to side fashion anastomosis | The novel side to side fashion anastomosis |
| --- | --- | --- |
| Technique difference | ● Cut distal outflow of the donor STA.<br>● End to side STA-MCA anastomosis. | ● Maintain distal outflow of the donor STA.<br>● Side to side STA-MCA anastomosis. |
| Advantages | ● Simple.<br>● Familiar by most neurosurgeons.<br>● Can choose any suitable PSCA in the surgical field as the recipient artery. | ● Self-regulating blood flow input based on patients' blood flow requirement.<br>● Accelerated EDAS.<br>● Has the potential to arouse all scalp arteries as donor sources for revascularization through the anastomotic site.<br>● Once CHS or watershed shift phenomenon occurred, the symptoms are relatively mild with short duration time.<br>● No scalp problem and interrupt of preexisting collaterals. |
| Disadvantages | ● Patients may passively accept mandatory blood flow input lack of self-regulating mechanism.<br>● Once CHS or watershed shift phenomenon occurred, the symptoms are relatively severe with long duration time. | ● Relatively unfamiliar by neurosurgeons.<br>● Somewhat technically limited.<br>● Can only choose the PSCAs near to the donor artery as the recipient artery. |

STA = superficial temporal artery; MCA= middle cerebral artery; PSCA = parasylvian cortical artery; EDAS = encephalo-duro-arterio-synangiosis; CHS = cerebral hyperperfusion syndrome.

### Limitations

The present study had several limitations. First, recipient selection is limited by the length and location of the donor artery. However, in this cohort, only one planned S-S anastomosis finally changed to E-S pattern due to no available recipient. On this condition, we can select E-S anastomosis even pure EDAS as a back-up in practice. Second, distal STA steal phenomenon may be a concern, so the patient and donor artery selection should be strict to avoid an inappropriate anastomosis. Third, this is a retrospective study in a single center and the number of patients is limited, further prospective controlled trials would be conducted to make S-S fashion can be widely used.

## Conclusions

Our study demonstrated that the S-S bypass is available and effective in the treatment for MMD patients. Compared to the traditional E-S bypass, S-S bypass can archive the similar perfusion and clinical improvement, but the milder manifestation and shorter duration time of postoperative CHS symptoms. Moreover, S-S bypass has the potential to arouse all scalp arteries as donor sources for revascularization through the intact distal STA branch by a flow self-regulating fashion. These novel principles make S-S bypass a new possibility for the routine treatment of MMD.

## Data Availability

All data produced in the present study are available upon reasonable request to the authors

## Acknowledgments

This study was partly supported by two projects of the National Natural Science Foundation of China (81671157 and 8217052644).

